# Leukopenia of Asymptomatic COVID-19 Infections under 18 Years Old in Recovery Stage

**DOI:** 10.1101/2020.05.21.20074682

**Authors:** Wei Zhang, Youshu Yuan, Zhengqiao Yang, Jinxia Fu, Yun Zhang, Ming Ma, Weidong Wu, Hourong Zhou

## Abstract

**Objectives:** In December, 2019, a type of novel coronavirus which was designated novel coronavirus 2019 (2019-nCoV) by World Health Organization (WHO) occurred in Wuhan, Hubei, China. The epidemiological and clinical characteristics of those patients under 18 years old in the recovery stage are limited. To compare the difference of epidemiological and clinical characteristics of COVID-19 involving 25 patients under 18 years old in recovery stage between confirmed and asymptomatic infections.

**Methods:** Retrospective, single-center cohort study of COVID-19 involving 25 patients under 18 years old in the recovery stage at Guizhou Provincial Staff Hospital in Guiyang, China, from January 29, to March 31, 2020; final date of follow-up was April 22. Epidemiological, demographic, clinical, laboratory, radiological, and treatment data were collected and analyzed. Epidemiological and clinical characteristics of confirmed COVID-19 infections and asymptomatic infections were compared.

**Results:** Among the 25 COVID infections under 18 years old, 16 (64%) were mild or moderate confirmed cases, and 9 (36%) were asymptomatic. The shortest treatment period was 6 days, the longest 26 days, and the average treatment period 14 days. Four cases (44.4%) had visited Wuhan or had a living story in the city. There were 9 (100%) asymptomatic cases were familial cluster outbreak, with an average infection number was 6 cases among all families. The number of asymptomatic COVID-19 infections with leukopenia were significantly more than confirmed cases (*p*=0.04).

**Conclusions:** Leukopenia mostly occurred in asymptomatic COVID-19 infections under 18 years old compared with the confirmed patients.

**Key Point:** In this single-center case series involving 25 cases under 18 years old with COVID-19 infections, leukopenia mostly occurred in asymptomatic infections.

## Background

Since December 2019, a type of novel coronavirus which was designated novel coronavirus 2019 (2019-nCoV) by World Health Organization (WHO) occurred in Wuhan, Hubei, China,^1^ and then it spread to the whole country and even the whole world^2,3^. On February 11, 2020, the WHO announced a new name for the epidemic disease caused by 2019-nCoV: 2019 Coronavirus Disease (COVID-19). As for the virus itself, the International Committee on Taxonomy of Viruses (ICTV) has renamed the previously provisionally named 2019-nCoV as severe acute respiratory syndrome coronaviruse-2, SARS-CoV-2.^4^ The origin of SARS-CoV-2 has not yet been explored, and early studies have found that wild animals such as pangolin were potential intermediate hosts of virus^5,6^, which means that the virus was carried by wild animals at the beginning, and now it was the first time to invade human beings. Meanwhile, it also means that human’s cognition of COVID-19 was not complete, and it was still in the process of exploration. China was the first country to experience COVID-19, and most of the early reported cases indicated middle-aged and elderly people or who existed diabetes, hypertension and cardiovascular diseases were common, children were few^7-10^. However, because of the immature immune system of children and with the spread of the epidemic and the number of children cases increases gradually, which should be paid more attention to children’s group.^11-13^ We aim to compare and analyze the epidemiological characteristics and physiological test data of asymptomatic patients and symptomatic group, explored whether there were significant differences between the two groups, so as to provide a theoretical basis for later clinical diagnosis and related studies.

## Methods

### Design

This is a retrospective, single-center cohort study of COVID-19 involving 25 patients under 18 years old from Guizhou Provincial Staff Hospital in the recovery period. CONSORT (Consolidated Standards of Reporting Trials) Flow Diagram of this study was unveiled in Figure 1. This study was retrospectively registered at the Chinese Clinical Trial Register (CCTR number: ChiCTR2000032458, registered 28 April 2020). URL: http://www.chictr.org.cn/edit.aspx?pid=52779&htm=4. The protocol of this study was designed by our team in March 15, 2020.

**Figure 1.**
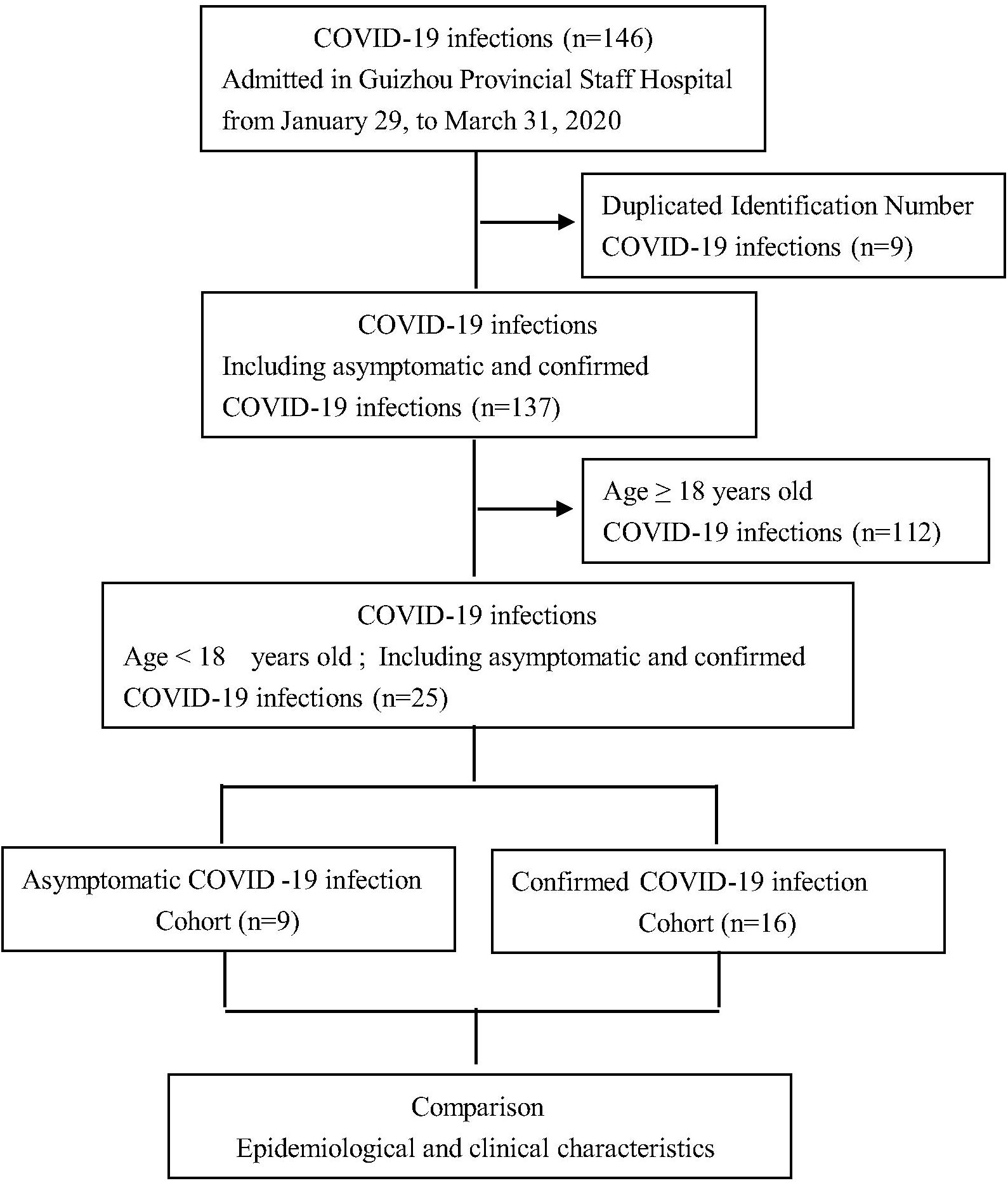
CONSORT Flow Diagram of This Study.

### Setting

A tertial hospital of COVID-19 medical center in Guizhou Province.

### Inclusion criteria

All asymptomatic and confirmed patients under 18 years old with COVID-19. All data were obtained from the records of a hospital information system (HIS) in Guizhou Provincial Staff Hospital.

### Exclusion criteria

All asymptomatic and confirmed patients of age ≥18 years old.

### Intervention

No.

### Main outcomes and measures

Epidemiological, demographic, clinical, laboratory, radiological, and treatment data were collected and analyzed. Epidemiological and clinical characteristics of confirmed COVID-19 infections and asymptomatic infections were compared.

### Study Definition

#### Classification

The standard for case classification in this study was based on COVID-19 diagnosis and treatment scheme, 1st to 7th edition. According to the national standards, mild and moderate patients were included in the symptomatic group in this study, while asymptomatic patients were included in the control group.

#### Epidemic focus

Since 2019-nCoV first appeared in Wuhan, Hubei province, and then spread to the whole country and even the whole world, most of the existing studies regard Wuhan or other areas of Hubei province as the Epidemic focus area.

#### Possible exposure time

That is the maximum probability to infection with COVID-19.

#### Incubation period

Theoretically, the incubation period is between onset infection and onset symptom. However, because the specific time of morbidity is affected by a variety of factors and cannot be accurately measured, we think the potential exposure time to the time that be allowed to get into hospital as the incubation period.

#### Treatment cycle

The number of days of treatment cycle were actually counted in this study, which refers to the time from the first diagnosis to the definite detection show the patient was cured, excluding the observation time transferred to the centralized isolation hospital.

#### Statistic Analyze

EXCEL software was used to arrange information, then choose suitable data imported into SPSS 26.0 for analysis. First of all, the method of mean sequence was used to process the missing values of the data. Considering the sample size was less than 40, Fisher’s Exact method was used to analyze the difference of class variable between the two groups. Independent sample T test was used for quantitative variables which conform to normal distribution among two groups. And non-parametric test of independent samples was used for variable data that not conform to normal distribution. All practices serve one purpose that is to compare the differences in epidemiological characteristics and physiological detection data between the two groups.

## Results

### CONSORT Flow Diagram (Figure 1)

In this research, the whole cohort encompassed 137 cases with COVID-19 infections including adults and children, as well as asymptomatic and confirmed COVID-19 infections. Twenty-five cases of age under 18 years old were divided into asymptomatic COVID-19 infections cohort and confirmed COVID-19 infection cohort, we respectively compared the epidemiological and clinical characteristics of the two groups.

### Basic epidemiological characteristics Disease type

Studies have shown that from the current cases who were sent to hospital to receive treatment. COVID-19 was less severe in patients under 18 years old, and most of were mild or moderate.^14^ However, there have been reports of critical illness in children at present^15^. Among the 25 cases in this study, 16 (64%, 16/25) had appeared infection symptoms, and 9 (36%, 9/25) had no symptoms. It should be pointed out that asymptomatic patients had no clinical performance, but it does not mean that they belong to patients with a mild degree of infection. The severity of infection should be determined according to the duration of virus activity in the body^16^. Moreover, asymptomatic patients are infectious and can still cause a certain range of transmission.^17^

### Gender and age composition

Among asymptomatic patients in the study, 5 (55.6%) cases of the patients were male. In terms of age, the youngest patient was only more than 5 months old, the oldest patient was 16 years old, the average age was 8.27 years old and the median age was 7 years old (inter-quartile range, 6 to 10). While 16 symptomatic patients made up of 10 (62.5%) male. From age structure to see, the youngest patient was an infant who only more than 2 months old, and the oldest patient was 17 years old, the average age was 9.6 years old, and the median age was 11 years ((inter-quartile range, 3.75 to 14.25). There was no significant difference in gender and age between the two groups (p=0.53 and p=0.34, respectively).

### Incubation

In pneumonia cases infected by SARS-CoV-2 in patients under 18 years old, the incubation is as short as one day and as long as 14 days. In this study, the mean sequence method was used to process the data about 8 patients’ incubation absence. The median incubation period for symptomatic patients was 10 days (inter-quartile range, 7 to 10), and 15 patients (93.75%, 15/16) had an incubation period of less than two weeks.

### Treatment circle

Previous studies have shown that the average treatment period of light and normal types was not more than two weeks. Collectible statistics show that the shortest period of asymptomatic patients was 6 days, the longest treatment period was 26 days, and the average treatment period was 14 days. For people who have symptoms the shortest treatment cycle was 4 days, the longest treatment cycle was 20 days, and the average treatment cycle was 12 days. The data showed that the average treatment circle of 9 asymptomatic patients was longer than two weeks and which was longer than symptomatic patients. However, there was no significant difference between the two groups in the treatment cycle.

### History of epidemiology

According to the existing epidemiological data, people who had visited Wuhan or had a living story in the city or other areas of Hubei province were those with high incidence of COVID-19, and close family contact was the key cause cluster outbreak in children. Among the asymptomatic patients, 4 (44.4%) had visited Wuhan or had a living story in the city or other areas of Hubei province. All the patients had familial cluster outbreak, and the average familial infection number was 6. As to symptomatic patients, 9 (56.3%) had visited Wuhan or had a living story in the city or other areas of Hubei province. Fourteen patients (87.5%) had familial clustering, and the familial average infection number was 5. By and large, there was no significant difference between the two groups in the three characteristics.

### Laboratory inspection

Among the asymptomatic patients in the recovery stage, 5 patients had decreased white blood cell count, 2 patients (22.2%) had increased lymphocyte absolute value, 3 patients had increased platelet count, and 4 patients had decreased neutrophils. Blood sedimentation was normal in all patients. Only 1 case (6.2%) of confirmed patients had decreased white blood cell count and 2 cases (12.5%) had increased in white blood cell count. The absolute value of lymphocytes was increased in 3 (18.75%) cases. 6 (37.5%) patients’ platelet count had risen. Neutrophils were decreased in 3 cases (18.75%). There were significant differences in Leukopenia between the two groups (*p*=0.04), but no significant differences in other laboratory test items.

### Chest computerized tomography (CT)

All the 25 patients underwent chest CT scan after getting into the hospital. Only 1 (11.11%) case of the asymptomatic patients had the characteristics of pulmonary infection (Figure 2), ground glass shadow change respectively located beside the right main bronchus in Panel A and dorsal inferior lobe of right lung in Panel B; Inflammatory consolidation shadows located in the dorsal inferior lobe of right lung in Panel C and D. Eight patients (88.89%, 8/9) had no distinct changes. Five (31.25%, 5/16) patients with symptomatic infection, CT indicated lesion change, and 11 (68.75%) patients had no change. The *p* value on CT suggested that there was no significant difference between the two groups. The CT of symptomatic patients did not change in accordance with the existing studies that there are a few patients with positive RT-PCR of throat swab test and a history of clinical characteristics and exposure in Hubei province, but there is no abnormal phenomenon in chest CT examination. Although real-time reverse-transcription–polymerase-chain-reaction (RT-PCR) is the gold standard for the detection of COVID-19, it presents a high false positive rate due to the influence of sample sampling and clinical medication. In view of chest CT is easy to perform, Similarly, it cannot achieve the complete accuracy of the detection results. Therefore, in order to ensure the accuracy of test results, both are often used conjunctively to measure COVID-19.

## Discussion

The occurrence of the 2019-nCoV in Hubei province aroused the attention of the provincial committee and government in Guizhou province, and then leading organization quickly formulates appropriate prevention and control measures. Guizhou province had the least COVID-19 patients in southwest China except Tibet autonomous region through joint effort. Since the first confirmed case of COVID-19 appeared in Guizhou province on January 13, 2020, the public health department actively executes joint prevention and control work, and launched the first-level response to public health emergencies on January 24, 2020. Until April 1, 2020, a total of 147 COVID-19 patients were confirmed, including 1 case imported from abroad, and 26 cases were children, accounting for 17.81% (26/146). In this study, there were 25 cases of children, have a proportion as high as 96.15% (25/26) of the cases in Guizhou province.

The reason that patients’ incubation period exist missing data is that 12 (48%, 12/25) children’s age at 10 years old or younger, due to the young did not have the ability to accurately describe onset time of their symptoms or even no memory about onset symptom. Another reasonable explanation is that asymptomatic patients have no obvious symptoms in the early stage of infection, so they are not aware of the fact of infection. In addition, the average treatment period among 9 asymptomatic patients was more than two weeks and longer than 2 days for symptomatic patients, and 3 (33.33%) patients’ treatment period was more than 3 weeks. The reason is that the long incubation period of asymptomatic patients made the virus survive in the body for a long time, and the degree of virus invasion was severe, so it needs to undergo a longer treatment cycle.

The literature has proved that in the routine blood test of COVID-19 patients, the occurrence of white blood cells or lymphocyte count decreased were common phenomenon^9,18^, and the number of white blood cells in both mild and moderate types is mostly decreased. In this study, 5 asymptomatic patients’ white blood cells had decreased, while only 1 symptomatic patient’s (6.2%) white blood cells had decreased and 2 (12.5%) cases had increased. There was a significant difference in white blood cells between the two groups (*p*=0.04). The reasonable explanation was that although the 5 asymptomatic patients had no clinical performance, they were infected with SARS-CoV-2 was a pre-exist fact. The erosion of the virus caused excessive destruction of white blood cells, so the number of white blood cells in peripheral blood had decreased. However, the presence of white blood cell count increased in patients with symptomatic was indicative that the aggravation of the patient’s condition. As for the significant difference in the white blood cell count between the two groups, it was considered that the difference in the immune system and the ability to resist the virus in the two groups.

### Advantages and disadvantages of this study

There are several advantages in our study. Firstly, this is the first study to use clinical inspection results and epidemiological survey data about COVID-19 in patients under 18 years old during the recovery stage. Secondly, the nature of our study was a retrospectively analysis that compared symptomatic with asymptomatic patients to draw conclusions. Thirdly, it is the first time that leukopenia is found mostly in asymptomatic patients under 18 years old with COVID-19.

Certainly, there are some disadvantages and shortages in the research. For example, the sample size of this study was a total of 25 patients, which was small and lacked credibility. Additionally, in view of the small number of COVID-19 patients in Guizhou province, the sample size of patients under 18 years old with COVID-19 in this study was small, so our conclusion will be verified by more studies in the future.

## Conclusions

In this study, most of patients under 18 years old infected by SARS-CoV-2 were mild or moderate type. Leukopenia mostly occurred in asymptomatic patients under 18 years old with COVID-19 than in those with confirmed infections.

## Declarations Ethics approval and consent to participate

We stated that written consent to participate was obtained from the parents or guardians of the minors who are under the age of 16, while patients aged 16-18 were obtained from themselves. This study was approved by the Biomedical Ethics Committee of Affiliated Hospital of Zunyi Medical University.

## Consent to publish

Not Applicable.

## Data Availability

We stated that all the data and materials were true and available in the study.

http://www.chictr.org.cn/edit.aspx?pid=52779&htm=4

## Availability of data and materials

We stated that all the data and materials were true and available in the study.

## Patent Data

These patients have not been reported in any other submission by you or anyone else.

## Authors’ contributions

ZW had full access to all data in the present study and accepts responsibility for data management and accuracy of the data analyses. Study concept and design: ZW, and ZHR. Acquisition and interpretation of data: FJX, Z Y and YZQ. Drafting of the manuscript: YYS, YZQ and FJX. Critical revision of the manuscript for important intellectual content: WWD and MM. Administrative, technical, or material support: ZW and ZHR. Study supervision: ZW and WWD. All authors agree to submission of the final version of this manuscript. ZW is the study guarantor.

## Acknowledgements

The authors thank Ping Lu, Min Yao, Yang Hong, Jinhua Wu, et.al. gave us selfish helps on the process of database construction of COVID-19 in this study.

## Authors’ Information

Youshu Yuan: Guizhou University School of Medicine, Guiyang, Guizhou, 550025, China.; Zhengqiao Yang: Guizhou Provincial Staff Hospital, Guiyang, Guizhou, 550025, China.; Jinxia Fu: Guizhou Provincial Staff Hospital, Guiyang, Guizhou, 550025, China.; Yun Zhang: Guizhou Provincial Staff Hospital, Guiyang, Guizhou, 550025, China.; Ming Ma: Guizhou Provincial Staff Hospital, Guiyang, Guizhou, 550025, China.; Weidong Wu: Guizhou Normal College, Guiyang, Guizhou, 550025, China.

## Figure Legends

Figure 2. Images Changes of a Boy with Asymptomatic COVID-19 Infection on Admission

